# Understanding and Usefulness of Effect Size and Certainty of Evidence: A Cross-sectional Survey of Evidence-Based Practice Competencies Among Registered Dietitians

**DOI:** 10.64898/2026.06.19.26356093

**Authors:** Nirjhar R. Ghosh, Elizabeth P. Neale, Rosa K. Hand, Geoff D.C. Ball, Emily Riddle, Kevin C. Klatt, Aaron Riviere, Bradley C. Johnston

## Abstract

**Introduction:** Understanding of absolute and relative estimates (i.e., effect size), and certainty of evidence corresponding to those estimates, is a fundamental evidence-based practice competency to promote informed clinical decision-making. While research has been conducted in the medical profession, there is no published research on these competencies in the nutrition and dietetics profession.

**Methods:** Among registered dietitians, our main objectives were to assess (1) their understanding and perceived usefulness of three absolute and two relative estimate approaches to assess effect size, (2) their perceived usefulness of certainty of evidence, and (3) factors influencing their understanding and perceived usefulness. We conducted a web-based, cross-sectional survey among dietitians recruited from the Academy of Nutrition and Dietetics (United States). Participants received effect estimates based on hypothetical dietary interventions vs. usual diet for reducing myocardial infarction risk.

**Results:** Of the 11,050 dietitians who received the survey link, 210 participated (2.0% response rate), and only completers (n=114) were included in the analysis. Participants demonstrated a similar understanding of the relative (27.6%) and absolute (27.5%) estimates, with Risk Difference (30.7% correct responses) being the best understood approach and Number Needed to Treat (24.6%) being the least. The understanding of five approaches was not different than random guessing (p>0.05). While perceived usefulness scores were similar between five approaches, they were highest when data was presented as Relative Risk [mean (SD): 4.82 (1.50)]. Dietitians rated the usefulness of certainty of evidence favorably [mean (SD): 5.07 (1.83), on a 7-point scale), and no factors were associated with correct understanding.

**Conclusion:** Dietitians may have limited understanding of how to interpret effect sizes, a finding consistent with surveys of other health professionals. To optimize informed decision-making between dietitians and clients, dietetic programs and continuing education platforms should consider additional training on interpreting effect sizes and certainty of evidence for effect sizes.

## INTRODUCTION

Evidence-based clinical decision-making in nutrition and dietetics involves asking structured clinical questions, systematically finding the best available research evidence and applying the evidence within the practice context, while considering clinical expertise and the values and preferences of patients to achieve favorable health outcomes [1,2]. The Academy of Nutrition and Dietetics’ (AND) 2024 standards of practice emphasize that registered dietitians should base nutrition interventions on the best available evidence and guidelines [3]. The Academy’s Code of Ethics further highlights the importance of evidence-based decisions, integrating patient values and practitioner expertise [4]. Similarly, accreditation standards from Canada’s Partnership for Dietetic Education and Practice, Dietitians Australia’s National Competency Standards for Dietitians and the British Dietetic Association stress equipping registered dietitians with evidence-based approaches to practice [5–7].

Typically, the best available evidence is derived from up to date systematic and comprehensive summaries of the research evidence, such as systematic reviews with meta-analyses (SRMAs), particularly when multiple studies exist [8,9]. Systematic reviews with (or without) meta-analyses are widely recognized as the fundamental building blocks of evidence-based practice (EBP), as they help prevent healthcare practitioners from being unduly influenced by personal biases, which can occur when clinicians are exposed to unrepresentative or low-quality evidence [10]. As with most health care professionals, registered dietitians (henceforth referred to as dietitians) are encouraged to rely on summary estimates from SRMAs to guide their clinical decisions [11]. To make rational decisions, dietitians must be competent in interpreting the results from SRMAs, in particular the absolute estimates for the outcomes that matter most to patients [12]. To present data for dichotomous outcomes such as mortality, myocardial infarction (fatal or non-fatal) or hospitalization, SRMAs commonly use relative estimates to express the difference in outcomes between intervention and control groups (also known as magnitude (size) of treatment/exposure effect or effect size) as a ratio (e.g., relative risk, odds ratio), and less commonly, the absolute estimates to optimally quantify the actual effect size (e.g., risk difference, number needed to treat) [13,14]. Understanding both types of estimates are considered fundamental competencies in EBP for health care professionals [12]. Systematic reviews with meta-analyses also report the certainty of evidence for each health outcome, which strengthens the confidence one can have in making recommendations to individual clients or for the purpose of practice guidelines [15,16]. It is crucial to consider the certainty of evidence alongside effect size in clinical decision-making; however, a recent systematic review of EBP competencies identified major gaps in the literature regarding empirical evidence on the correct understanding of absolute and relative estimates and interpreting the certainty of evidence for these estimates among dietitians [2]. In this study, we aimed to address these gaps by surveying dietitians from the United States.

## MATERIALS AND METHODS

### Study objectives

Our primary objective was to assess dietitians’ correct understanding of five approaches of effect estimate. We tested their understanding of the magnitude (size) of three absolute estimates: risk difference (RD) together with absolute risk reduction (ARR), risk in treatment/exposure vs. control groups and number needed to treat (NNT), and two relative estimates: relative risk (RR) together with relative risk reduction (RRR) and odds ratio (OR) together with relative odds reduction (ROR). We also evaluated dietitians’ perceived usefulness for each of the five approaches (e.g., absolute and relative estimates) for understanding the effect size. Additionally, we assessed their views on the usefulness of presenting certainty of evidence alongside the effect estimates for informing clinical decisions. We used the certainty ratings of very low, low, moderate and high from the Grading of Recommendations Assessment, Development and Evaluation (GRADE) approach because they are commonly used in SRMAs [16]. Our secondary objective was to identify factors associated with correct understanding of the absolute and relative estimates and those influencing dietitians’ perceived usefulness of these estimates. We hypothesized that dietitians would demonstrate better understanding and perceive greater usefulness for absolute estimates (i.e., RD/ARR, NNT, risk in treatment/exposure vs. control groups) compared to relative estimates (i.e., RR/RRR, OR/ROR). Additionally, we hypothesized that dietitians would find it useful to have certainty of evidence presented alongside these estimates for clinical decision-making.

### Study design and sample

We conducted a web-based, cross-sectional survey among dietitians randomly selected from the AND registry (United States) using the ‘Qualtrics’ platform (confidentiality policy: https://www.qualtrics.com/privacy-statement). Eligibility criteria included current involvement in clinical practice (part-time or full-time), teaching or research, proficiency in English, ability to provide consent and use online survey platforms. The sample size was determined based on an estimated 28% proportion of correct responses from prior studies on interpreting statistical formats by physicians [17]. To achieve a 5% margin of error with 95% confidence, the estimated sample size was 308, which was deemed sufficient to assess variations in understanding and perceived usefulness across absolute and relative estimates. Our study is reported in accordance with the Consensus-based checklist for Reporting of Survey Studies guideline [18], and the survey was deemed exempt by the Texas A&M University Internal Review Board (IRB2023-0909M).

### Survey distribution

The survey was distributed via the AND registry and remained accessible from May 13, 2024 to July 31, 2024. Recruitment emails were sent twice by the Academy’s web marketing team to 10,000 randomly selected dietitians. We also targeted approximately 1,050 additional dietitians through two dietetic practice groups from the AND (Nutrition Educators of Health Professionals, Research). Because individuals may belong to both the registry and practice groups, the number of unique individuals who received the invitation cannot be determined. The survey was anonymous and needed to be completed in one session. To ensure unique participation, we used Qualtrics’ built-in tools for reCAPTCHA and bot detection and collected IP addresses, which were not linked with participant responses. This prevented individuals from participating more than once, even if they received the survey invitation through multiple channels (AND registry, dietetic practice groups). Random assignment ensured even distribution between both versions.

### Questionnaire development

We developed a questionnaire tailored for dietitians, drawing from a similar survey that assessed physicians’ understanding of effect estimates [17]. The questionnaire began with demographic and background questions. We then presented a clinical scenario based on a hypothetical SRMA of randomized controlled trials. The SRMA examined the relationship of five theoretically different dietary interventions vs. usual diet on the risk of developing a fatal or non-fatal (with serious consequences) myocardial infarction over 5 years of follow-up [19]. We chose “theoretically” different diets, without naming the diet interventions, to avoid participants’ preconceived ideas about certain diets. So that study participants could focus on effect estimates rather than concerns about systematic errors (bias) or random errors (precision), we described the estimates as having a low risk of bias with precise effect estimates (i.e., narrow confidence intervals and very low p-values [p<0.001]). Participants were also asked not to focus on issues like cost or inconveniences associated with dietary changes. Finally, they were instructed to answer without utilizing any external resources such as search engines (e.g., Google) or Generative AI.

Following the scenario, participants answered questions on their understanding of five effect estimate approaches and perceived usefulness of the approaches for understanding the effect size, with one question per estimate (totaling ten questions). Participants also rated the usefulness of having certainty of evidence ratings alongside effect estimates (one question), and were provided with an open-ended question to explain their reasoning for the rating.

To assess understanding, we used multiple-choice questions, where the options for classifying the effect size included: trivial (probably not important), small (probably important), moderate (surely important) and large (very important) [20]. Each question had only one correct answer. To rate their perceived usefulness for each of the five approaches and certainty of evidence, participants used a 7-point Likert-type scale (1 = not useful at all, 7 = extremely useful). To examine whether different approaches influenced their understanding of effect size, we developed two versions of the survey with large and trivial effect size. Upon accessing the survey link, participants were randomly assigned to either a ‘large’ effect scenario (ARR of 6.0%) or a ‘trivial’ effect scenario (ARR of 0.2%). For each version, based on two similar surveys conducted among medical professionals [17,21], we determined trivial and large effect sizes for each of our approaches based on consultations involving three methodologists and one biostatistician (**Appendix A**). Both versions underwent expert review through user testing with five dietitians from academic institutions in the USA, Canada and Australia. Their feedback on clarity and formatting informed iterative revisions, resulting in the final version. Face validity was assessed to ensure the questionnaire measured its intended purpose, based on the subjective judgment of the expert team, who represented the target survey participants (i.e., registered dietitians). The complete survey is provided in **Appendix B**.

### Data analysis

We used descriptive statistics to summarize participants’ sociodemographic characteristics (e.g., biological sex, ethnicity), professional attributes (e.g., work settings, years of research experience) and educational background (e.g., highest degree obtained, training in EBP). For categorical outcomes, such as participants’ understanding of the effect size for five approaches, we calculated the frequency and percentage of correct answers along with corresponding 95% confidence intervals (CIs). For continuous outcomes, such as participants’ perceived usefulness of five approaches and certainty of evidence, we reported means and standard deviations (SD) along with corresponding 95% CIs.

We used Cochran’s Q test and repeated-measures ANOVA to determine differences in understanding and perceived usefulness, respectively, across five approaches. To compare perceived usefulness between trivial and large effects, we used independent samples t-tests. We explored the association of seven *a priori* factors with correct understanding and perceived usefulness: primary work setting, years involved in research, job hours as a clinician, years of clinical experience, highest degree obtained, training in health research (e.g., biostatistics, epidemiology) and training in EBP. We used chi-square test or Fisher’s Exact test for understanding, and one-way ANOVA and independent samples t-tests for perceived usefulness. We categorized open-ended responses on the perceived usefulness of certainty of evidence as either supportive or critical and included example quotations for each category. Post-hoc, we conducted binomial tests to assess if correct understanding differed from chance alone, and Wilcoxon signed-rank tests to compare understanding and paired-samples t-tests to compare perceived usefulness between absolute and relative approaches across trivial and large effect sizes.

We set the significance threshold at α = 0.05 for all statistical tests and conducted analyses using IBM SPSS Statistics for Windows, version 29 (IBM Corp., Armonk, N.Y., USA). Only fully completed survey questionnaires were included in the analysis, while participants with missing data, due to unanswered questions or ambiguous responses, were excluded.

## RESULTS

Approximately 11,050 dietitians received the survey link and 210 accessed it (2.0% response rate), who were then randomly assigned to either the trivial or large effect group. Of those, 114 (54.3%) fully completed the questionnaire (trivial group, n=55; large group, n=59) with an average completion time of 15 minutes. **Appendix Figure 1** shows the participant flow chart. Among 114 completers, 89.5% were female, 6.1% identified as Hispanic or Latino, 87.7% were white and 40.4% held a master’s degree. Almost 38% completed master’s-level epidemiology or biostatistics courses and almost 38% enrolled in EBP courses during undergraduate or graduate training. Nearly all respondents (94.8%) had provided direct nutrition care to individuals or groups at some point in their careers, with clinical experience ranging from one year to over 21 years. At the time of the survey, 53.5% worked in clinical settings (inpatient, outpatient, ambulatory or long-term care), 14% were involved in research with or without teaching, and 10.5% were involved in private practice (**Table 1**). The participants’ demographic characteristics were compared with the 2024 Compensation and Benefits Survey [22], revealing that our sample aligned closely with nationally representative data. For instance, the 2024 survey reported that 92% of respondents were female, 6% identified as Hispanic or Latino, 90% were White, 52% held a master’s degree, 55% worked in clinical settings (inpatient, outpatient, ambulatory or long-term care) and 5% were involved in private practice.

**Table 1:** Characteristics of dietitians who completed the survey.

| **Characteristic** | **No (%) of participants** |
| --- | --- |
| Female | 102 (89.5%) |
| Male | 7 (6.1%) |
| Non-Hispanic or Latino | 107 (93.9%) |
| Hispanic or Latino | 7 (6.1%) |
| White | 100 (87.7%) |
| Black or African American | 4 (3.5%) |
| Asian | 3 (2.6%) |
| Multiracial | 4 (3.5%) |
| Specialty certification as an RD |  |
| None | 82 (71.9%) |
| Advanced Diabetes Management | 4 (3.5%) |
| Certified Nutrition Support Dietitian/Clinician | 11 (9.6%) |
| Current primary work setting |  |
| Clinical Nutrition | 61 (53.5%) |
| Social Services/Public Health | 7 (6.1%) |
| Primarily Research (PhD/postdoc/academic positions) | 8 (7.0%) |
| Primarily Teaching (College/University) | 7 (6.1%) |
| Both Research & Teaching | 8 (7.0%) |
| Private Practice | 12 (10.5%) |
| Government | 2 (1.8%) |
| Highest degree in Nutrition or Dietetics |  |
| Baccalaureate Degree | 48 (42.1%) |
| Master's Degree | 46 (40.4%) |
| Doctoral Degree | 14 (12.3%) |
| Level of epidemiology/biostatistics courses completed |  |
| Baccalaureate level | 12 (10.5%) |
| Master's level | 43 (37.7%) |
| Doctoral level | 15 (13.2%) |
| None | 41 (36%) |
| Enrolled in an EBP course during undergrad or grad training | 43 (37.7%) |
| Participated in any EBP-focused continuing education | 20 (17.5%) |
| Years involved in research |  |
| None | 65 (57.0%) |
| 1 to 5 years | 27 (23.7%) |
| 6 to 10 years | 5 (4.4%) |
| 11 to 15 years | 5 (4.4%) |
| 16 to 20 years | 6 (5.3%) |
| 21 years or more | 6 (5.3%) |
| Amount of time working in a clinical setting |  |
| Full time ($\geq 35$ hours/week) | 40 (35.1%) |
| Part time (under 35 hours/week) | 27 (23.7%) |
| Per diem (as needed) | 18 (15.8%) |
| Not involved with clinical practice | 29 (25.4%) |
| Years spent providing clinical care to individuals/groups |  |
| None | 6 (5.3%) |
| 1 to 5 years | 45 (39.5%) |
| 6 to 10 years | 17 (14.9%) |
| 11 to 15 years | 13 (11.4%) |
| 16 to 20 years | 9 (7.9%) |
| 21 years or more | 24 (21.1%) |

### Understanding and perceived usefulness of five approaches

Overall, participants demonstrated a similar understanding of the relative (27.6%) and absolute (27.5%) estimate approaches (p = 0.78). Their understanding was highest for RD/ARR (30.7% correct responses), closely followed by OR/ROR (29.8%). Risk in treatment vs. control groups was the third most understood approach (27.2%), followed by RR/RRR (25.4%), and the lowest understanding was for NNT (24.6%) (**Table 2**). However, the binomial test showed that understanding of all five approaches was not different than random guessing (p-values>0.05, **Table 2**). Dietitians’ perceived usefulness was highest for RR/RRR [mean (SD): 4.82 (1.50)] for understanding the effect size, followed by OR/ROR [4.74 (1.50)], risk in treatment vs. control groups, RD/ARR [4.61 (1.62)], and NNT [4.33 (1.82)] (**Table 2**).

**Table 2.**
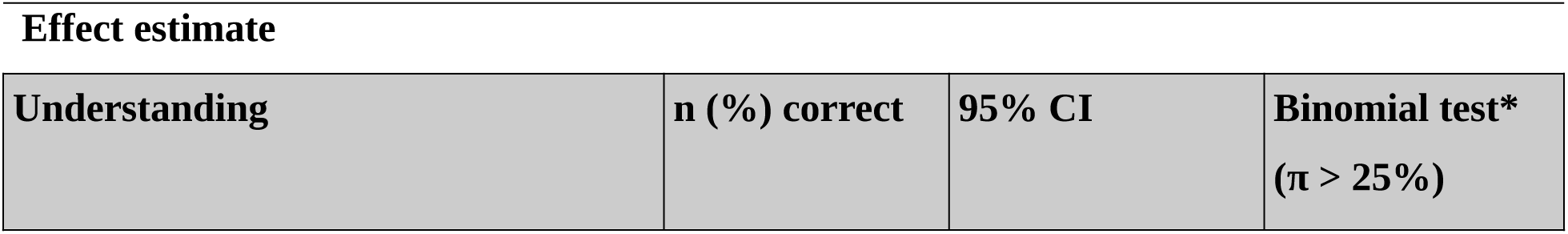
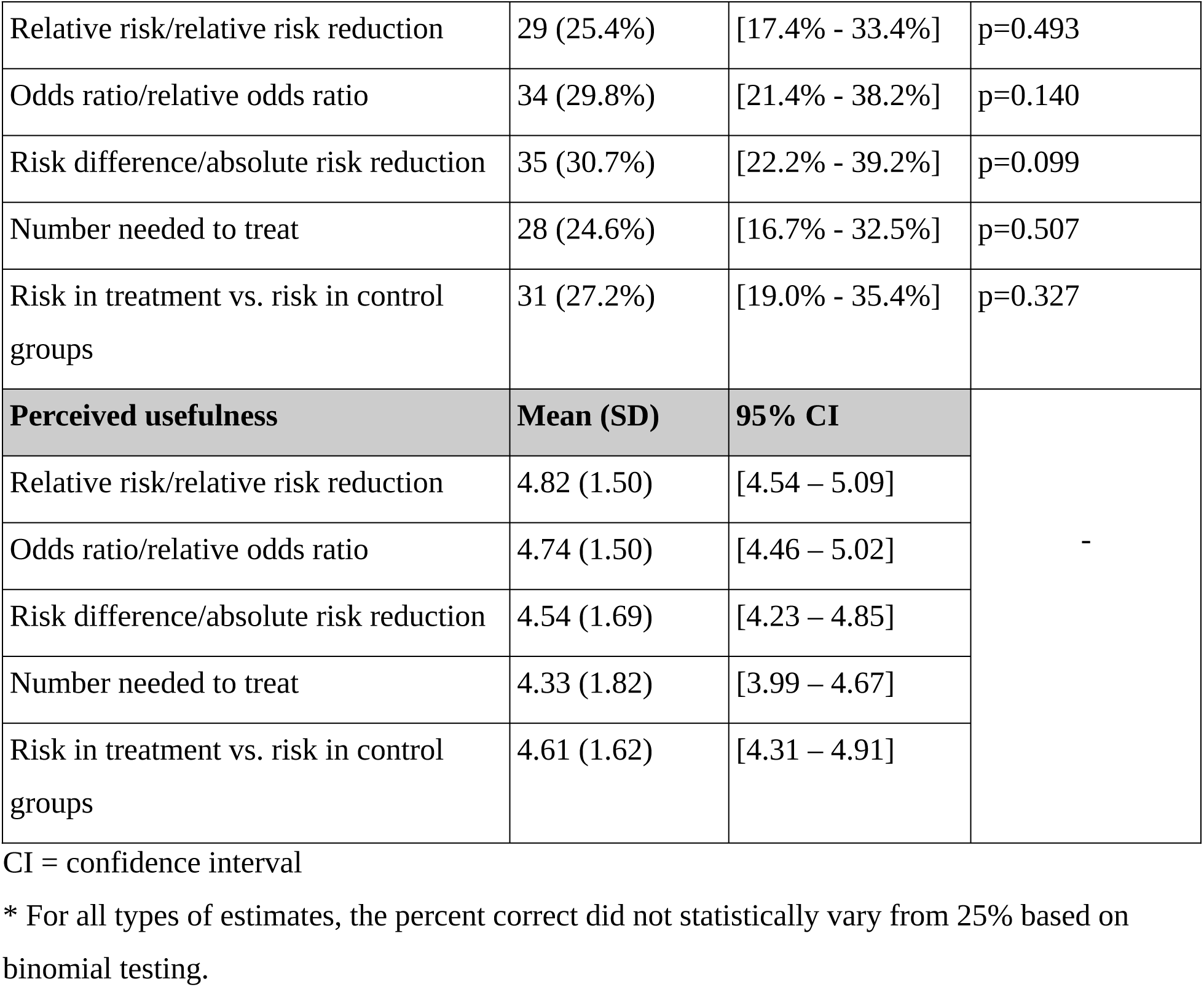
Correct understanding of treatment/exposure effect size from five effect estimate approaches.

### Understanding and perceived usefulness across trivial and large effect size

Dietitians’ understanding of trivial effects was better for absolute estimate approaches. For instance, the highest understood approach was RD/ARR (60%), followed by NNT (45.5%) and risk in treatment vs. control groups (43.6%). In contrast, for large effects, dietitians showed a higher understanding of relative estimates. For instance, the most understood approach was OR/ROR (45.8%), followed by RR/RRR (42.4%). In terms of perceived usefulness, for trivial effect size, dietitians rated risk in treatment/exposure vs. control groups the highest [4.44 (1.73)], followed by OR/ROR [4.36 (1.58)]. For large effect size, they rated RR/RRR the highest [5.25 (1.28)], followed by OR/ROR [5.08 (1.34)]. **Table 3** illustrates these comparisons across understanding and perceived usefulness for our five approaches.

**Table 3.** Comparison between absolute vs relative estimates under trivial and large treatment/exposure effect size.

| Analysis | Type of measure |  |  |
| --- | --- | --- | --- |
|  | Absolute estimates | Relative estimates | Comparison between types |
| **Entire sample, n = 114** |  |  |  |
| Understanding<sup>1</sup> [score 0-1] | 0.27 (0.35) | 0.28 (0.41) | p=0.783 |
| Perceived usefulness<sup>2</sup> [score 1-7] | 4.50 (1.48) | 4.78 (1.43) | **p=0.012** |
| **Trivial effects, n = 55** |  |  |  |
| Understanding<sup>1</sup> [score 0-1] | 0.50 (0.38) | 0.10 (0.28) | **p<0.001** |
| Perceived usefulness<sup>2</sup> [score 1-7] | 4.33 (1.61) | 4.35 (1.49) | p=0.870 |
| **Large effects, n = 59** |  |  |  |
| Understanding<sup>1</sup> [score 0-1] | 0.07 (0.14) | 0.44 (0.45) | **p<0.001** |
| Perceived usefulness<sup>2</sup> [score 1-7] | 4.65 (1.34) | 5.17 (1.25) | **p=0.002** |

Reported values are mean (SD)
<sup>1</sup> understanding score is a proportion of correct measures ranging from 0 (none correct) to 1 (all correct)
<sup>2</sup> perceived usefulness is assessed on 7-point Likert scale (1 = not useful at all ... 7 = extremely useful)

### Factors associated with understanding and perceived usefulness of five approaches

No factors were associated with understanding; however, respondents with EBP training demonstrated higher perceived usefulness for RR/RRR (p=0.004) and RD/ARR (p=0.010). **Appendix Tables 1** and **2** present the relationships between various factors and respondents’ understanding and perceived usefulness.

### Perceived usefulness of certainty of evidence

Participants rated their perceived usefulness of having certainty of evidence alongside effect estimates for clinical decision-making favorably [mean (SD): 5.07 (1.83), on a 7-point scale]. Of the 17 participants who provided open-ended responses, most were supportive of the GRADE certainty approach, while a few described it as arbitrary and subjective. Key attributes derived from these responses, along with example quotations, are presented in **Table 4**.

**Table 4:**
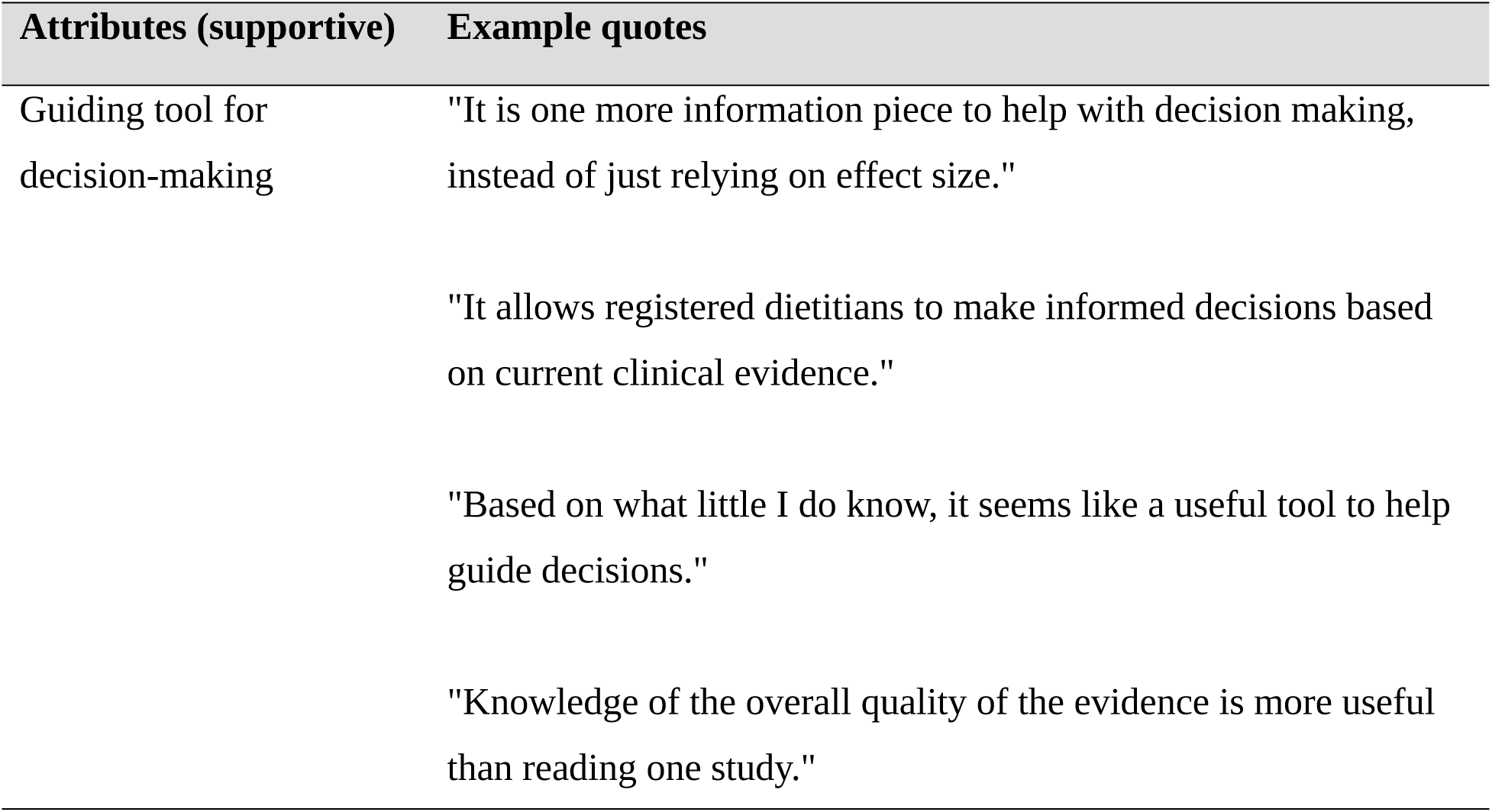
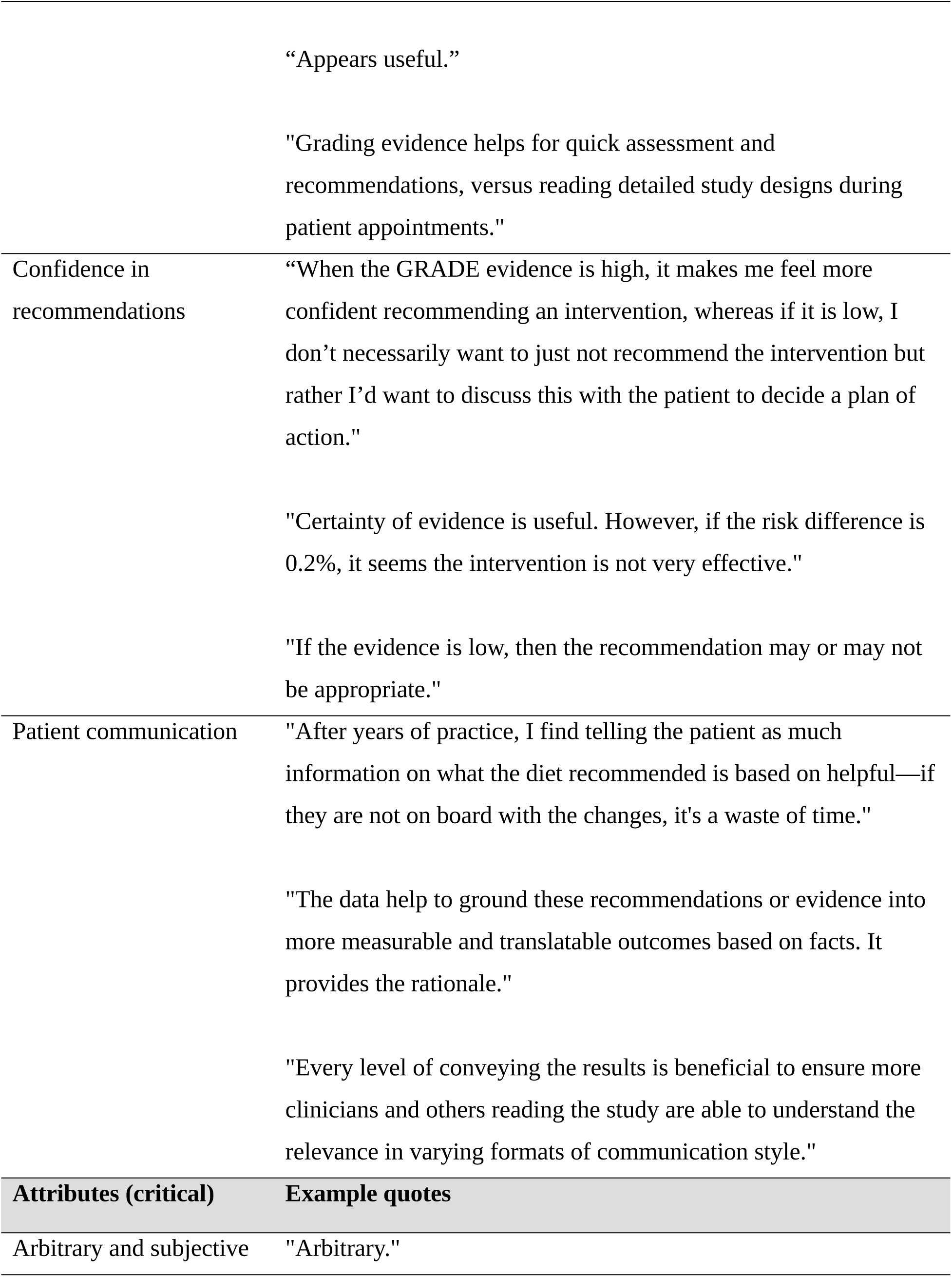
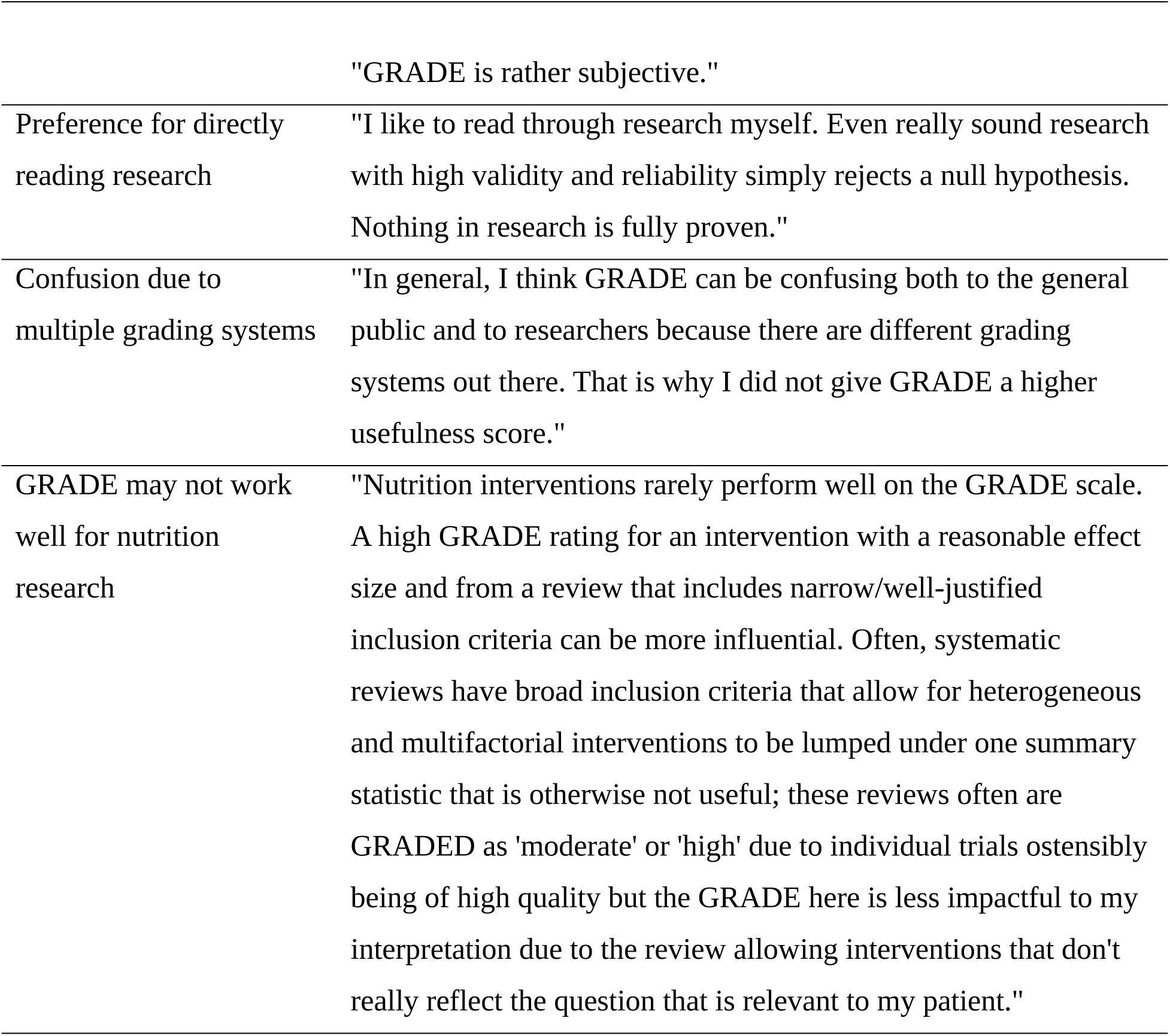
Quotations illustrating support or critics of certainty of evidence.

| Attributes (supportive) | Example quotes |
| --- | --- |
| Guiding tool for decision-making | "It is one more information piece to help with decision making, instead of just relying on effect size." |
|  | "It allows registered dietitians to make informed decisions based on current clinical evidence." |
|  | "Based on what little I do know, it seems like a useful tool to help guide decisions." |
|  | "Knowledge of the overall quality of the evidence is more useful than reading one study." |

|  | “Appears useful.”<br>"Grading evidence helps for quick assessment and recommendations, versus reading detailed study designs during patient appointments." |
| --- | --- |
| Confidence in recommendations | “When the GRADE evidence is high, it makes me feel more confident recommending an intervention, whereas if it is low, I don’t necessarily want to just not recommend the intervention but rather I’d want to discuss this with the patient to decide a plan of action."<br>"Certainty of evidence is useful. However, if the risk difference is 0.2%, it seems the intervention is not very effective."<br>"If the evidence is low, then the recommendation may or may not be appropriate." |
| Patient communication | "After years of practice, I find telling the patient as much information on what the diet recommended is based on helpful—if they are not on board with the changes, it's a waste of time."<br>"The data help to ground these recommendations or evidence into more measurable and translatable outcomes based on facts. It provides the rationale."<br>"Every level of conveying the results is beneficial to ensure more clinicians and others reading the study are able to understand the relevance in varying formats of communication style." |
| Attributes (critical) | Example quotes |
| Arbitrary and subjective | "Arbitrary." |
|  | "GRADE is rather subjective." |
| Preference for directly reading research | "I like to read through research myself. Even really sound research with high validity and reliability simply rejects a null hypothesis. Nothing in research is fully proven." |
| Confusion due to multiple grading systems | "In general, I think GRADE can be confusing both to the general public and to researchers because there are different grading systems out there. That is why I did not give GRADE a higher usefulness score." |
| GRADE may not work well for nutrition research | "Nutrition interventions rarely perform well on the GRADE scale. A high GRADE rating for an intervention with a reasonable effect size and from a review that includes narrow/well-justified inclusion criteria can be more influential. Often, systematic reviews have broad inclusion criteria that allow for heterogeneous and multifactorial interventions to be lumped under one summary statistic that is otherwise not useful; these reviews often are GRADED as 'moderate' or 'high' due to individual trials ostensibly being of high quality but the GRADE here is less impactful to my interpretation due to the review allowing interventions that don't really reflect the question that is relevant to my patient." |

## DISCUSSION

In our survey of dietitians, their understanding was generally consistent across the five approaches of effect estimate, with no statistically significant difference between relative and absolute estimates. Although not statistically significant, understanding was highest for RD/ARR (an absolute estimate). Previous studies among healthcare professionals, including physicians, nurses, pharmacists and dentists, also showed highest understanding for absolute estimates [17,21]. Our binomial test indicated that the correct understanding observed for each of the five approaches may have resulted from chance rather than dietitians’ true knowledge of the topic. This raises concerns about dietitians’ ability to accurately interpret effect estimates regardless of how they are presented (i.e., relative vs. absolute). Respondents viewed relative estimates as more useful, ranking RR/RRR as the most helpful and NNT, an absolute estimate, as the least. This disconnect, where RR/RRR ranked fourth in comprehension but highest in perceived usefulness, suggests that dietitians may be relying on familiarity rather than comprehension when judging usefulness. They may be more familiar with relative estimates, which are commonly used in health science literature, compared to absolute estimates [13,23]. Indeed, while absolute estimates for dichotomous outcomes are essential to informed and intuitive decision-making for clinicians and patients, they are unfortunately rarely provided in nutrition research [24–26]. Respondents who received EBP training rated higher usefulness for RD/ARR and RR/RRR, suggesting that EBP training may improve appreciation of different approaches for communicating effect size. In general, the inclusion of certainty of evidence alongside effect estimates was perceived as useful for more informed clinical decision-making. This finding is particularly important given that informed clinical decisions require not only an understanding of the estimated effect size, but also an understanding of the certainty associated with that effect.

### Strengths

First, this is the first survey to assess dietitians’ objective knowledge with respect to interpreting effect sizes for both absolute and relative estimates, and to explore dietitians’ perceived usefulness of having certainty of evidence ratings presented alongside effect estimates. Second, given that <5% of SRMAs in nutrition report certainty of evidence to accompany effect estimates [25,26] and no prior research has examined dietitians’ views on the usefulness of certainty of evidence in clinical decision-making, we addressed an important gap related to communicating nutrition science results. The perceived usefulness and comfort with certainty of evidence ratings based on the GRADE approach may be limited by or relate to some ongoing controversy around GRADE’s utility versus other alternatives (e.g. NutriGrade) [30–32]. We argue that GRADE is well-suited and readily adaptable to the field of nutrition and dietetics. Criticisms of GRADE are readily circumvented by appropriate engagement between evidence synthesis and nutrition experts and consideration of discipline-specific factors affecting the certainty of evidence, such the importance of effect modification by non-placebo comparators and habitual intakes/nutrient status as well as the accuracy of exposure assessment through both biomarkers and memory-based dietary intake assessment [33,34]. Alternatives to GRADE do not present unique benefits for assessing the certainty of evidence in nutrition but do result in standards in nutrition deviating from virtually all other areas of biomedicine and healthcare. Dietetic educators involved in the training of future dietitians as well as those developing continuing education materials should enthusiastically embrace GRADE to ensure the profession of dietetics meets modern standards of EBP and can readily interface with systematic evidence reviews and authoritative guidance, which includes those published by the AND, Cochrane and the Dietary Reference Intakes [35,36]. Third, the demographic characteristics of our sample were similar to a nationally representative data [22], supporting its representativeness of the broader dietetic population.

### Limitations

Our study has limitations as well. First, despite efforts to increase our sample size by including two dietetic practice groups from the AND, our sample was small, requiring caution when interpreting the findings. Second, we used effect size categories based on GRADE guidance [20], and the term ‘trivial’ may be less familiar to dietitians than categories used in other approaches, such as Cohen’s *d* (small, moderate, large). This may have affected participants’ understanding of effect size. Third, in our original questionnaire, we included questions assessing dietitians’ confidence in their understanding of the effect size. However, their confidence varied in different directions and did not correspond to our usefulness data, as with past surveys of medical professionals [17,21]. Given the findings did not yield logical results, we assumed that their confidence was low and that they randomly chose their confidence ratings, thus we have relegated the results on confidence to our appendices (**Appendix Table 3**). The disconnect in dietitians’ confidence and perceived usefulness may be best explained by a lack of foundational training and knowledge in interpreting effect sizes and certainty of evidence. Two of the authors teach in nutrition dietetic programs and through continuing education platforms for dietitians (BCJ, KCK), and in all instances we have noticed that our teaching on effect size and certainty of evidence was both novel and highly appreciated by learners.

### Implication for clinical practice

Although dietitians best understood RD/ARR (30.7%) and least understood NNT (24.6%), their understanding was no different than randomly guessing the answers. This low understanding of effect size regardless of the effect estimate approach, along with the absence of meaningful differences in perceived usefulness between absolute and relative estimates, indicates an important limitation in foundational knowledge needed to support EBP. Participants’ feedback at the end of the survey further highlighted this, with comments like, “I am not certain about the statistics being analyzed,” and “I am unfamiliar with number [needed] to treat and risk difference and risk in both groups, so it was difficult to rank these terms [for usefulness].” These challenges highlight the need for comprehensive, standardized EBP training that would help dietitians understand effect estimates and categorize the effect size as, for example, trivial, small, medium or large [2,20]. Furthermore, our findings highlight the necessity of dietetics training on certainty of evidence for effect estimates. For example, some dietitians recognized its usefulness but were unsure how to apply it. One respondent noted, “[GRADE] appears useful, but I am not certain how to use it with patients,” while another stated, “I need more training in this.” Integrating training into nutrition curricula at undergraduate, graduate and continuing education levels and clinical workflows will help bridge this gap [27]. The need for further training in competencies related to interpreting effect size and certainty of evidence is underscored by the scale of the profession, 113,266 registered dietitians and 11,414 dietetics students and interns in the United States as of 2025, and growing emphasis on shared clinical decision-making [28,29].

### Implications for research

Future studies should aim for a larger, more diverse sample, ideally across multiple countries, to ensure statistical power and generalizability. Further, expanding recruitment to include dietetic students and interns would provide insights into whether dietetic training programs adequately prepare future dietitians for interpreting effect sizes when presented as both absolute and relative estimates, as well as certainty of evidence ratings.

## CONCLUSION

Our survey showed that dietitians had a similar understanding of five approaches of effect estimate with no good evidence of participants’ ability to discern effect size, whether presented as an absolute (e.g., ARR, NNT) or a relative estimate (e.g., RR/RRR, OR/ROR). That is, the correct understanding of effect size was not better than random chance alone. Dietitians rated the usefulness of presenting certainty of evidence favorably. Our findings underscore the need for more research on dietitians’ competencies specific to effect size, and the likely need for additional training and education on interpreting effect size and certainty of evidence.

## Data Availability

All data produced in the present work are contained in the manuscript.

## Acknowledgment

We would like to thank Anton Svendrovski (StatsHelp.ca) for his assistance with the statistical analysis.

## Author contributions

Bradley C. Johnston and Nirjhar R. Ghosh conceived the study, designed it and developed the questionnaire. Nirjhar R. Ghosh collected the data, interpreted the results and drafted the first version of the manuscript. All authors critically revised the manuscript and contributed to the final version.

## Data Availability

The Consensus-based checklist for Reporting of Survey Studies is available upon request.

