## Supplementary file for "Understanding and Usefulness of Effect Size and Certainty of Evidence: A Cross-sectional Survey of Evidence-Based Practice Competencies Among Registered Dietitians"

**Supplementary Materials**

### Appendix A: Summary and rationale for large and trivial treatment/exposure effects

Large and trivial effects are listed in the table below, followed by the rationale for these effects based on GRADE guidance and the Users' Guides to the Medical Literature. Our outcome of interest was myocardial infarction, with the large versus trivial effects anchored on a small but important improvement in the risk of myocardial infarction (fatal and non-fatal), and patient important outcome.

| **Approach** | **Large effect from dietary intervention** | **Large effect interpretation** | **Trivial effect from dietary intervention** | **Trivial effect interpretation** |
|---|---|---|---|---|
| Relative risk/Relative risk reduction | RR of 0.50, RRR of 50% | Large relative reduction in risk, very important to patients | RR of 0.90, RRR of 10% | Small relative reduction in risk, probably not important to patients |
| Odds ratio/Relative odds reduction | OR of 0.47, ROR of 53% | Large reduction in odds, very important to patients | OR of 0.89, ROR of 11% | Small reduction in odds, probably not important to patients |
| Risk difference/Absolute risk reduction | RD/ARR of 0.06 (6%) | Large absolute reduction in risk, very important to patients | RD/ARR of 0.002 (0.2%) | Small absolute reduction in risk, probably not important to patients |
| Number needed to treat | NNT 17 | Low NNT (greater benefit), very important to patients | NNT 500 | High NNT (less benefit), probably not important to patients |
| Risk in treatment vs. control groups | 6% vs. 12% (i.e., RD of 6%) | Large difference in risk between groups, very important to patients | 1.8% vs. 2% (i.e., RD of 0.2%) | Small difference in risk between groups, probably not important to patients |

#### Rationale:

#### Relative Risk (RR) and Relative Risk Reduction (RRR)

These estimates are based on GRADE guidelines 9 (Rating up the certainty of evidence) [1]. The guideline states that for relative risk, the magnitude of effect may be considered large if the RR is 0.5 to 0.2 (RRR of 50% to 80%) if and when no plausible confounders have been ruled out. Based on this guidance, we assumed that:

- Large (very important) effect: RR of 0.5 to 0.2 (RRR of 50% to 80%),
- Moderate (surely important) effect: RR of 0.6 to 0.7 (RRR of 40% to 30%),
- Small (probably important) effect: RR of 0.8 to 0.88 (RRR of 20% to 12%),
- Trivial (probably not important) effect: RR of ≤0.89 (≤11%).

#### Odds Ratio (OR) and Relative Odds Reduction (ROR)

These estimates are also based on GRADE guidelines 9 [1]. The guideline notes that when baseline risk is low (<20%), the odds ratio and relative risk are very similar, and one can apply the relative risk criteria to estimate the effect size for odds ratio. Since our baseline risk was <20% in both scenarios (trivial and large), we applied the same RR/RRR thresholds to OR/ROR:

- Large (very important) effect: OR of 0.5 to 0.2 (ROR of 50% to 80%),
- Moderate (surely important) effect: OR of 0.6 to 0.7 (ROR of 40% to 30%),
- Small (probably important) effect: OR of 0.8 to 0.88 (ROR of 20% to 12%),
- Trivial (probably not important) effect: OR of ≤0.89 (ROR of ≤11%).

#### Risk Difference (RD) and Absolute Risk Reduction (ARR)

These estimates are based on GRADE guidelines 6 (Rating the quality of evidence—imprecision) [2]. The guideline states that an ARR of ≥1% (RD of ≥0.01) is considered important to patients. We therefore defined:

- Large (very important) effect: ARR ≥5% (RD ≥0.05),
- Moderate (surely important) effect: ARR of 3% to 4% (RD of 0.03 to 0.04),
- Small (probably important) effect: ARR of 1% to 2% (RD of 0.01 to 0.02),
- Trivial (probably not important) effect: ARR <1% (RD <0.01).

#### Number Needed to Treat (NNT)

The NNT is the inverse of the risk difference (calculated as: 1 ÷ RD) [3]. Thus, we used the criteria for RD to define the effect size for NNT:

- Large (very important) effect: NNT ≤20 (RD ≥0.05),
- Moderate (surely important) effect: NNT 25–33 (RD 0.03–0.04),
- Small (probably important) effect: NNT 50–100 (RD 0.01–0.02),
- Trivial (probably not important) effect: NNT >100 (RD <0.01).

#### Risk in Treatment vs. Control Groups

This is calculated by subtracting the experimental event rate from the control event rate (an alternative expression of RD/ARR). Therefore, following the assumption of ARR/RD categories, we defined:

- Large (very important) effect: ARR of ≥5% (RD of ≥0.05) in treatment vs. control groups,
- Moderate (surely important) effect: ARR of 3% to 4% (RD of 0.03 to 0.04) in treatment vs. control groups,

- Small (probably important) effect: ARR of 1% to 2% (RD of 0.01 to 0.02) in treatment vs. control groups,
- Trivial (probably not important) effect: ARR of <1% (RD of <0.01) for risk in treatment vs. control groups.

Therefore, in our case, for a large effect we considered the risk in treatment (6%) vs. control groups (12%) [ARR of 6% (RD of 0.06)], calculated by subtracting experimental event rate (n=60) from control event rate (n=120). For a trivial effect, we considered the risk in treatment (1.8%) vs. control groups (2%) [ARR of 0.2% (RD of 0.002)], calculated by subtracting experimental event rate (n=18) from control event rate (n=20).

### Appendix B: Survey questionnaire for trivial & large effects with correct answers

*Trivial effect size version*

Please answer the following questions to the best of your knowledge.

What is your gender identity?

- O Woman
- O Man
- O Transgender
- O Non-binary
- O Other
- O Prefer not to answer

What is your ethnic origin?

- O Hispanic or Latino
- O Not Hispanic or Latino

What is your race?

- O White
- O Black or African American
- O American Indian or Alaska Native
- O Asian
- O Native Hawaiian or other Pacific Islander
- O Multiracial

- ○ Other

Do you currently hold any specialty certification as a Registered Dietitian? [select one or more]

- ☐ Board Certified Advanced Diabetes Management (BC-ADM)/Certified Diabetes Educator (CDCES)
- ☐ Board Certified Specialist in Oncology Nutrition (CSO)
- ☐ Board Certified Specialist in Gerontological Nutrition (CSG)
- ☐ Board Certified Specialist in Pediatric Nutrition (CSP)
- ☐ Board Certified Specialist in Renal Nutrition (CSR)
- ☐ Board Certified Specialist in Sports Dietetics (CSSD)
- ☐ Board Certified Specialist in Pediatric Critical Care Nutrition (CSPCC)
- ☐ Board Certified Specialist in Obesity and Weight Management (CSOWM)
- ☐ Certified Nutrition Support Dietitian/Clinician (CNSD/C)
- ☐ Clinical Lipid Specialist (CLS)
- ☐ None
- ☐ Other

If selected 'Other', please specify:

________________________________________

What is your (current) primary work setting?

- O Clinical Nutrition – Inpatient, Outpatient, Long-term care, Ambulatory
- O Social Services/Public Health
- O Food Service/Management
- O Primarily Research (PhD and postdoctoral positions, academic positions, jobs in research and development sectors)
- O Primarily Teaching (College/University)
- O Both Research & Teaching
- O Government
- O Private Practice
- O Industry
- O Other

If selected 'Primarily Research' or 'both Research & Teaching', how many years have you been involved in research?

- O 1 to 5 years
- O 6 to 10 years
- O 11 to 15 years
- O 16 to 20 years
- O 21 years or more
- O None

As a Registered Dietitian, what amount of time do you work in a clinical setting?

- O Full time (≥ 35 hours/week)
- O Part time (under 35 hours/week)
- O Per diem (as needed)
- O I am not involved with clinical practice
- O Other
- O

If selected 'Other', please specify:

________________________________________________________________

How many years have you spent in clinical practice providing direct nutrition care to individuals or groups?

- O 1 to 5 years
- O 6 to 10 years
- O 11 to 15 years
- O 16 to 20 years
- O 21 years or more
- O None

What is your highest degree (held or in progress)?

- O Baccalaureate Degree
- O Master's Degree
- O Doctoral Degree
- O Master's Degree in Progress
- O Doctoral Degree in Progress

What is your field of study for the highest degree (held or in progress)?

________________________________________________________________

What is your highest degree in Nutrition or Dietetics (held or in progress)?

- O Baccalaureate Degree
- O Master's Degree
- O Doctoral Degree
- O Master's Degree in Progress
- O Doctoral Degree in Progress
- O Not applicable

Year of completion (or expected completion year) for the highest degree in Nutrition or Dietetics:

________________________________________________________________

Did you complete (or are you completing) any courses in epidemiology or biostatistics that provided training in data presentation methods (e.g., relative risk, risk difference), certainty/strength of evidence?

- O Yes
- O No

If selected 'Yes', what level of courses did you complete (or are you completing)?

- O Baccalaureate level
- O Master's level
- O Doctoral level
- O Master's level in Progress
- O Doctoral level in Progress

Following 2 questions are about your previous training and experience with evidence-based practice (EBP). This section offers details about the definition of EBP and its corresponding competencies. We have adapted our EBP definition and competencies based on the Academy of Nutrition and Dietetics and the JAMA Users' Guides to the Medical Literature. These competencies fall under the foundational EBP domains: Ask, Acquire, Appraise, Interpret and Apply.

We define EBP as a procedure that promotes shared decision-making between clinicians and patients based on three fundamental principles: (i) Use of best available research evidence (e.g., best evidence would be high-quality up-to-date practice guidelines or systematic review of the totality of evidence, followed by single randomized clinical trials, then cohort studies, then case-control studies, etc.) (ii) Clinical or real-world experience (iii) Consideration of patients' values and preferences based on the evidence for potential benefits and harms of an intervention We define EBP competencies as knowledge, skills, attitudes and behaviors in: i) Formulating a structured clinical question ii) Finding the best available research evidence iii) Assessing the methodological quality of the best available research evidence iv) Assessing the study results (i.e., magnitude of treatment/exposure effects [often referred to as 'effect size'] and the precision of effects) for all potential benefits and harms v) Interpreting the certainty of evidence for all potential benefits and harms based on study results vi) Applying results to clinical care based on the generalizability of the evidence to one's patient, including the patient's values and preferences based on the evidence for potential benefits and harms of an intervention.

During your undergraduate or graduate training, did you enroll in an EBP course?

(Please respond 'Yes' only if you attended an EBP course that covered any of the EBP competencies stated above)

O Yes

O No

If selected 'Yes', please describe the length and emphasis of the course with respect to the EBP competencies.

[As a reminder, we define EBP competencies as knowledge, skills, attitudes and behaviors in: i) Formulating a structured clinical question, ii) Finding the best available research evidence, iii) Assessing the methodological quality of the best available research evidence, iv) Assessing the study results (i.e., magnitude of treatment/exposure effects [often referred to as 'effect size'] and the precision of effects) for all potential benefits and harms, v) Interpreting the certainty of evidence for all potential benefits and harms based on study results, vi) Applying results to clinical care based on the generalizability of the evidence to one's patient, including the patient's values and preferences based on the evidence for potential benefits and harms of an intervention.]

________________________________________________________________

Have you participated in any EBP-focused continuing professional education (CPE) or related seminars, webinars, workshops or conferences (e.g., Duke University Teaching and Leading EBP Workshop; Texas A&M Workshop on Evidence-Based Nutrition Practice, Case Western Evidence-Based Practice for Healthcare Professionals)?

O Yes

O No

If selected 'Yes', please describe the length and emphasis of the course with respect to the EBP competencies.

[As a reminder, we define EBP competencies as knowledge, skills, attitudes and behaviors in: i) Formulating a structured clinical question, ii) Finding the best available research evidence, iii) Assessing the methodological quality of the best available research evidence, iv) Assessing the study results (i.e., magnitude of treatment/exposure effects [often referred to as 'effect size'] and the precision of effects) for all potential benefits and harms, v) Interpreting the certainty of evidence for all potential benefits and harms based on study results, vi) Applying results to clinical care based on the generalizability of the evidence to one's patient, including the patient's values and preferences based on the evidence for potential benefits and harms of an intervention.]

---

UNDERSTANDING: The following part of the survey presents a clinical scenario informed by a hypothetical but high-quality, up-to-date systematic review with meta-analysis of randomized controlled trials with precise estimates (narrow 95% confidence intervals, p-value<0.001). Please use the scenario and the meta-analysis results to answer the following questions. We request you to focus solely on the data presentation methods (and your perceived interpretation of the effect size) and the certainty of evidence related to the effect size, and not on other potential outcomes or issues like cost or inconvenience associated with dietary changes. Please answer without utilizing any external resources (e.g., Google, ChatGPT).

CLINICAL SCENARIO: *A 50-year-old man with several risk factors for developing heart disease (e.g., family history of heart attack, smoking, sedentary lifestyle, unhealthy dietary habit) is referred to you for primary prevention of heart attack. He has a BMI of 30. Given his family history, the patient is interested in decreasing his risk of heart attack based on your best interpretation of the available dietary intervention evidence.*

Imagine you have done a thorough search in PubMed/Medline related to your client's condition. The best available evidence you found is a recent systematic review with meta-analysis of 20 randomized controlled trials. The review analyzed the effects of five different dietary interventions vs usual diet on the risk of developing a fatal or non-fatal (with serious consequences) heart attack over 5 years of follow-up. The review shows a Relative Risk of 0.90 (representing a Relative Risk Reduction of 10%), favoring dietary intervention A. It means if patients follow intervention A for a 5-year period, they have 0.9 times less risk of developing a heart attack compared to the patients who follow a usual diet. In other words, the intervention group has (on average) a 10% Relative Risk Reduction of developing a heart attack compared to those following a usual diet.

How would you interpret the effect size of a Relative Risk of 0.90 (or a Relative Risk Reduction of 10%) in this case?

- O Trivial, probably not important
- O Small, probably important
- O Moderate, surely important
- O Large, very important

How confident are you about your interpretation of the effect size for Relative Risk (or Relative Risk Reduction)?

| | 1 Not confident at all | 2 | 3 | 4 | 5 | 6 | 7 Extremely confident |
|---|---|---|---|---|---|---|---|
| Please use the Likert scale to answer this question. | O | O | O | O | O | O | O |

How useful do you find the presentation of a Relative Risk (or a Relative Risk Reduction) for understanding effect size?

| | 1 Not useful at all | 2 | 3 | 4 | 5 | 6 | 7 Extremely useful |
|---|---|---|---|---|---|---|---|
| Please use the Likert scale to answer this question. | O | O | O | O | O | O | O |

The review shows an Odds Ratio of 0.89 (representing a Relative Odds Reduction of 11%), favoring dietary intervention B. It means if patients follow intervention B for a 5-year period, the odds of developing a heart attack is 0.89 times less likely compared to the patients who follow a usual diet. In other words, the odds of developing a heart attack are 11% lower in the intervention group relative to the usual diet group. How would you interpret the effect size of an Odds Ratio of 0.89 (or a Relative Odds Reduction of 11%) in this case?

- O Trivial, probably not important
- O Small, probably important
- O Moderate, surely important
- O Large, very important

How confident are you about your interpretation of the effect size for Odds Ratio (or Relative Odds Reduction)?

| | 1 Not confident at all | 2 | 3 | 4 | 5 | 6 | 7 Extremely confident |
|---|---|---|---|---|---|---|---|
| Please use the Likert scale to answer this question. | O | O | O | O | O | O | O |

How useful do you find the presentation of an Odds Ratio (or a Relative Odds Reduction) for understanding effect size?

| | 1 Not useful at all | 2 | 3 | 4 | 5 | 6 | 7 Extremely useful |
|---|---|---|---|---|---|---|---|
| Please use the Likert scale to answer this question. | O | O | O | O | O | O | O |

The review shows a Risk Difference in heart attack of 0.002 (representing an Absolute Risk Reduction of 0.2%) for dietary intervention C. That means if 1000 patients follow intervention C for a 5-year period, 2 fewer patients will have a heart attack from the intervention group compared to the usual diet group. How would you interpret the effect size of a Risk Difference of 0.002 (or an Absolute Risk Reduction of 0.2%) in this case?

O Trivial, probably not important

O Small, probably important

O Moderate, surely important

O Large, very important

How confident are you about your interpretation of the effect size for Risk Difference (or Absolute Risk Reduction)?

| | 1 Not confident at all | 2 | 3 | 4 | 5 | 6 | 7 Extremely confident |
|---|---|---|---|---|---|---|---|
| Please use the Likert scale to answer this question. | O | O | O | O | O | O | O |

How useful do you find the presentation of a Risk Difference (or an Absolute Risk Reduction) for understanding effect size?

| | 1 Not useful at all | 2 | 3 | 4 | 5 | 6 | 7 Extremely useful |
|---|---|---|---|---|---|---|---|
| Please use the Likert scale to answer this question. | O | O | O | O | O | O | O |

The review shows that 500 patients must be treated with dietary intervention D for a 5-year period to prevent 1 additional heart attack. Here, 500 patients is the 'Number Needed to Treat'. How would you interpret the effect size of a Number Needed to Treat of 500?

O Trivial, probably not important

O Small, probably important

O Moderate, surely important

O Large, very important

How confident are you about your interpretation of the effect size for Number Needed to Treat?

| | 1 Not confident at all | 2 | 3 | 4 | 5 | 6 | 7 Extremely confident |
|---|---|---|---|---|---|---|---|
| Please use the Likert scale to answer this question. | O | O | O | O | O | O | O |

How useful do you find the presentation of a Number Needed to Treat for understanding effect size?

| | 1 Not useful at all | 2 | 3 | 4 | 5 | 6 | 7 Extremely useful |
|---|---|---|---|---|---|---|---|
| Please use the Likert scale to answer this question. | O | O | O | O | O | O | O |

The review shows that if 1000 patients follow dietary intervention E for a 5-year period, 18 will have a heart attack (here, the risk of having a heart attack from intervention E is 1.8%). Alternatively, if those 1000 patients follow a usual diet for a 5-year period, 20 will have a heart attack (here, the risk of having a heart attack from usual diet is 2%). How would you compare the risk between intervention group and usual diet group?

O Trivial, probably not important

O Small, probably important

O Moderate, surely important

O Large, very important

How confident are you about your interpretation of the effect size for risk in both groups?

| | 1 Not confident at all | 2 | 3 | 4 | 5 | 6 | 7 Extremely confident |
|---|---|---|---|---|---|---|---|
| Please use the Likert scale to answer this question. | O | O | O | O | O | O | O |

How useful do you find the presentation of risk in both groups for understanding effect size?

| | 1 Not useful at all | 2 | 3 | 4 | 5 | 6 | 7 Extremely useful |
|---|---|---|---|---|---|---|---|
| Please use the Likert scale to answer this question. | O | O | O | O | O | O | O |

Please drag the data presentation methods to rank them from Most useful to Least useful for understanding the effect size. 1 = Most useful; 5 = Least useful

_______ Relative risk/Relative risk reduction
_______ Odds ratio/Relative odds reduction
_______ Risk difference
_______ Number needed to treat
_______ Risk in both groups

Now, you have the best estimate of effect regarding dietary intervention C on the risk of having a heart attack, which is a 0.2% risk difference for a 5-year period. Suppose you also know the certainty of evidence for this estimate. In the review, certainty of evidence was determined using the GRADE framework (used by Academy of Nutrition and Dietetics).

Certainty of evidence refers to how confident we are in the 'certainty' of an intervention effect on the target outcome (e.g., heart attack). The GRADE certainty ratings are defined as:

a) Very low certainty: The true effect is probably markedly different from the estimated effect

b) Low certainty: The true effect might be markedly different from the estimated effect

c) Moderate certainty: The true effect is probably close to the estimated effect

d) High certainty: The authors have a lot of confidence that the true effect is similar to the estimated effect

If you are aware that the certainty of evidence for a Risk Difference of 0.2% is ‘High’, would you recommend intervention C for your patient?

- O Yes
- O No
- O I don't know
- O I will share the results with my patient to determine the answer

Continue to suppose the best estimate of effect (risk difference) is 0.2% for a 5-year period. However, if you are aware that the certainty of evidence is ‘Low’ for the estimate of effect, would you recommend intervention C for your patient?

- O Yes
- O No
- O I don't know
- O I will share the results with my patient to determine the answer

For clinical decision making, how useful do you find having GRADE certainty of evidence (e.g., low, high) presented together with data presentation methods (e.g., relative risk, risk difference) to express the effect size?

| | 1 Not useful at all | 2 | 3 | 4 | 5 | 6 | 7 Extremely useful |
|---|---|---|---|---|---|---|---|
| Please use the Likert scale to answer this question. | O | O | O | O | O | O | O |

Please provide any comments or reasoning for your answer to the previous question.

________________________________________________________________

### Understanding and Usefulness of Effect Size and Certainty of Evidence: A Cross-Sectional Survey of Evidence-Based Practice Competencies Among Registered Dietitians

*Large effect size version*

UNDERSTANDING: The following part of the survey presents a clinical scenario informed by a hypothetical but high-quality, up to date, systematic review with meta-analysis with precise estimates (narrow 95% confidence intervals, p-value<0.001). Please use the scenario and the meta-analysis results to answer the following questions. We request you to focus solely on the data presentation methods (and your perceived interpretation of the effect size) and the certainty of evidence related to the effect size, and not on other potential outcomes or issues like cost or inconvenience associated with dietary changes. Please answer without utilizing any external resources (e.g., Google, ChatGPT). CLINICAL SCENARIO: *A 50-year-old man with several risk factors for developing heart disease (e.g., family history of heart attack, smoking, sedentary lifestyle, unhealthy dietary habit) is referred to you for primary prevention of heart attack. He has a BMI of 30. Given his family history, the patient is interested in decreasing his risk of heart attack based on your best interpretation of the available dietary intervention evidence.* Imagine you have done a thorough search in PubMed/Medline related to your client's condition. The best available evidence you found is a recent systematic review with meta-analysis of 20 randomized controlled trials. The review analyzed the effects of five different dietary interventions vs usual diet on the risk of developing a fatal or non-fatal (with serious consequences) heart attack over 5 years of follow-up. The review shows a Relative Risk of 0.50 (representing a Relative Risk Reduction of 50%), favoring dietary intervention A. It means if patients follow intervention A for a 5-year period, they have 0.5 times less risk of developing a heart attack compared to the patients who follow a usual diet. In other words, the intervention group has (on average) a 50% Relative Risk Reduction of developing a heart attack compared to those following a usual diet. How would you interpret the effect size of a Relative Risk of 0.50 (or a Relative Risk Reduction of 50%) in this case?

- O Trivial, probably not important
- O Small, probably important
- O Moderate, surely important
- O Large, very important

How confident are you about your interpretation of the effect size for Relative Risk (or Relative Risk Reduction)?

| | 1 Not confident at all | 2 | 3 | 4 | 5 | 6 | 7 Extremely confident |
|---|---|---|---|---|---|---|---|
| Please use the Likert scale to answer this question. | O | O | O | O | O | O | O |

How useful do you find the presentation of a Relative Risk (or a Relative Risk Reduction) for understanding effect size?

| | 1 Not useful at all | 2 | 3 | 4 | 5 | 6 | 7 Extremely useful |
|---|---|---|---|---|---|---|---|
| Please use the Likert scale to answer this question. | O | O | O | O | O | O | O |

The review shows an Odds Ratio of 0.47 (representing a Relative Odds Reduction of 53%), favoring dietary intervention B. It means if patients follow intervention B for a 5-year period, the odds of developing a heart attack is 0.47 times less likely compared to the patients who follow a usual diet. In other words, the odds of developing a heart attack are 53% lower in the intervention B group relative to the usual diet group.    How would you interpret the effect size of an Odds Ratio of 0.47 (or a Relative Odds Reduction of 53%) in this case?

- O Trivial, probably not important
- O Small, probably important
- O Moderate, surely important
- O Large, very important

How confident are you about your interpretation of the effect size for Odds Ratio (or Relative Odds Reduction)?

| | 1 Not confident at all | 2 | 3 | 4 | 5 | 6 | 7 Extremely confident |
|---|---|---|---|---|---|---|---|
| Please use the Likert scale to answer this question. | O | O | O | O | O | O | O |

How useful do you find the presentation of an Odds Ratio (or a Relative Odds Reduction) for understanding effect size?

| | 1 Not useful at all | 2 | 3 | 4 | 5 | 6 | 7 Extremely useful |
|---|---|---|---|---|---|---|---|
| Please use the Likert scale to answer this question. | O | O | O | O | O | O | O |

The review shows a Risk Difference in heart attack of 0.06 (representing an Absolute Risk Reduction of 6%) for dietary intervention C. That means if 1000 patients follow intervention C for a 5-year period, 60 fewer patients will have a heart attack from the intervention group compared to the usual diet group. How would you interpret the effect size of a Risk Difference of 0.06 (or an Absolute Risk Reduction of 6%) in this case?

O Trivial, probably not important

O Small, probably important

O Moderate, surely important

O Large, very important

How confident are you about your interpretation of the effect size for Risk Difference (or Absolute Risk Reduction)?

| | 1 Not confident at all | 2 | 3 | 4 | 5 | 6 | 7 Extremely confident |
|---|---|---|---|---|---|---|---|
| Please use the Likert scale to answer this question. | O | O | O | O | O | O | O |

How useful do you find the presentation of a Risk Difference (or an Absolute Risk Reduction) for understanding effect size?

| | 1 Not useful at all | 2 | 3 | 4 | 5 | 6 | 7 Extremely useful |
|---|---|---|---|---|---|---|---|
| Please use the Likert scale to answer this question. | O | O | O | O | O | O | O |

The review shows that 17 patients must be treated with dietary intervention D for a 5-year period to prevent 1 additional heart attack. Here, 17 patients is the 'Number Needed to Treat'. How would you interpret the effect size of a Number Needed to Treat of 17?

O Trivial, probably not important

O Small, probably important

O Moderate, surely important

O Large, very important

How confident are you about your interpretation of the effect size for Number Needed to Treat?

| | 1 Not confident at all | 2 | 3 | 4 | 5 | 6 | 7 Extremely confident |
|---|---|---|---|---|---|---|---|
| Please use the Likert scale to answer this question. | O | O | O | O | O | O | O |

How useful do you find the presentation of a Number Needed to Treat for understanding effect size?

| | 1 Not useful at all | 2 | 3 | 4 | 5 | 6 | 7 Extremely useful |
|---|---|---|---|---|---|---|---|
| Please use the Likert scale to answer this question. | O | O | O | O | O | O | O |

The review shows that if 1000 patients follow dietary intervention E for a 5-year period, 60 will have a heart attack (here, the risk of having a heart attack from intervention E is 6%). Alternatively, if those 1000 patients follow a usual diet for a 5-year period, 120 will have a heart attack (here, the risk of having a heart attack from usual diet is 12%). How would you compare the risk between intervention group and usual diet group?

O Trivial, probably not important

O Small, probably important

O Moderate, surely important

O Large, very important

How confident are you about your interpretation of the effect size for risk in both groups?

| | 1 Not confident at all | 2 | 3 | 4 | 5 | 6 | 7 Extremely confident |
|---|---|---|---|---|---|---|---|
| Please use the Likert scale to answer this question. | O | O | O | O | O | O | O |

How useful do you find the presentation of risk in both groups for understanding effect size?

| | 1 Not useful at all | 2 | 3 | 4 | 5 | 6 | 7 Extremely useful |
|---|---|---|---|---|---|---|---|
| Please use the Likert scale to answer this question. | O | O | O | O | O | O | O |

Please drag the data presentation methods to rank them from Most useful to Least useful for understanding the effect size. 1 = Most useful; 5 = Least useful

______ Relative risk/Relative risk reduction
______ Odds ratio/Relative odds reduction
______ Risk difference
______ Number needed to treat
______ Risk in both groups

Now, you have the best estimate of effect regarding dietary intervention C on the risk of having a heart attack, which is a 6% risk difference for a 5-year period. Suppose you also know the certainty of evidence for this estimate. In the review, certainty of evidence was determined using the GRADE framework (used by Academy of Nutrition and Dietetics). Certainty of evidence refers to how confident we are in the 'certainty' of an intervention effect on the target outcome (e.g., heart attack). The GRADE certainty ratings are defined as: a) Very low certainty: The true effect is probably markedly different from the estimated effect b) Low certainty: The true effect might be markedly different from the estimated effect c) Moderate certainty: The true effect is probably close to the estimated effect d) High certainty: The authors have a lot of confidence that the true effect is similar to the estimated effect If you are aware that the certainty of evidence for a Risk Difference of 6% is ‘High’, would you recommend intervention C for your patient?

- O Yes
- O No
- O I don't know
- O I will share the results with my patient to determine the answer

Continue to suppose the best estimate of effect (risk difference) is 6% for a 5-year period. However, if you are aware that the certainty of evidence is ‘Low’ for the estimate of effect, would you recommend intervention C for your patient?

- O Yes
- O No
- O I don't know
- O I will share the results with my patient to determine the answer

For clinical decision making, how useful do you find having GRADE certainty of evidence (e.g., low, high) presented together with data presentation methods (e.g., relative risk, risk difference) to express the effect size?

| | 1 Not useful at all | 2 | 3 | 4 | 5 | 6 | 7 Extremely useful |
|---|---|---|---|---|---|---|---|
| Please use the Likert scale to answer this question. | O | O | O | O | O | O | O |

Please provide any comments or reasoning for your answer to the previous question.

________________________________________________________________

Thank you for taking the survey, we appreciate your time and contribution.

**Appendix Figure 1.** Participant flow chart

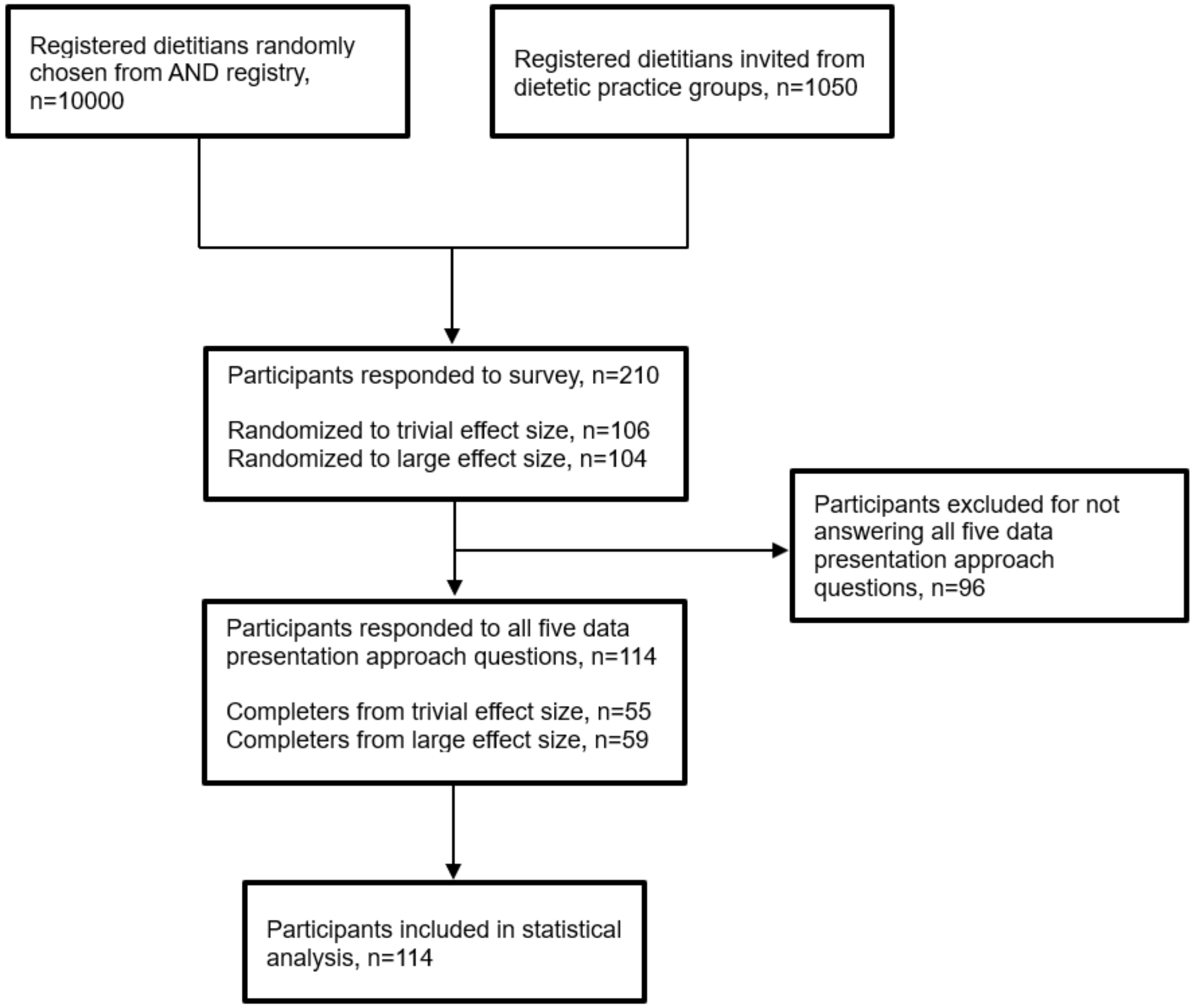

### Appendix Table 1. Factors associated with understanding of five approaches

| Variables | Approach | | | | |
|---|---|---|---|---|---|
| | **RR/RRR** | **OR/ROR** | **RD/ARR** | **NNT** | **Risk in treatment vs. control groups** |
| Primary work setting | $p = 0.916^{F}$ | $p = 0.995^{F}$ | $p = 0.233^{F}$ | $p = 0.078^{F}$ | $p = 0.560^{F}$ |
| Clinical Nutrition | 16 (26.2%) | 19 (31.1%) | 21 (34.4%) | 15 (24.6%) | 16 (26.2%) |
| Social Services/Public Health | 3 (42.9%) | 3 (42.9%) | 3 (42.9%) | 3 (42.9%) | 3 (42.9%) |
| Primarily Research | 2 (25.0%) | 2 (25.0%) | 1 (12.5%) | 0 (0%) | 1 (12.5%) |
| Primarily Teaching | 2 (28.6%) | 2 (28.6%) | 3 (42.9%) | 2 (28.6%) | 4 (57.1%) |
| Both Research & Teaching | 3 (37.5%) | 3 (37.5%) | 2 (25.0%) | 3 (37.5%) | 2 (25.0%) |
| Government | 0 (0%) | 0 (0%) | 0 (0%) | 1 (50.0%) | 0 (0%) |
| Private Practice | 2 (16.7%) | 3 (25.0%) | 1 (8.3%) | 1 (8.3%) | 2 (16.7%) |
| Industry | 0 (0%) | 0 (0%) | 2 (100%) | 2 (100%) | 1 (50.0%) |
| Other | 1 (14.3%) | 2 (28.6%) | 2 (28.6%) | 1 (14.3%) | 2 (28.6%) |
| Primary work setting | $p = 0.771^{F}$ | $p = 1.00^{F}$ | $p = 0.943^{F}$ | $p = 0.509^{F}$ | $p = 0.730^{F}$ |
| Clinical practice | 18 (24.7%) | 22 (30.1%) | 22 (30.1%) | 16 (21.9%) | 18 (24.7%) |
| Non-clinical practice | 10 (29.4%) | 10 (29.4%) | 11 (32.4%) | 11 (32.4%) | 11 (32.4%) |
| Other | 1 (14.3%) | 2 (28.6%) | 2 (28.6%) | 1 (14.3%) | 2 (28.6%) |
| Years involved in research | $p = 0.229^{F}$ | $p = 0.064^{F}$ | $p = 0.818^{F}$ | $p = 0.372^{F}$ | $p = 0.898^{F}$ |
| None | 14 (21.5%) | 20 (30.8%) | 22 (33.8%) | 14 (21.5%) | 20 (30.8%) |
| 1 to 5 years | 7 (25.9%) | 8 (29.6%) | 7 (25.9%) | 8 (29.6%) | 6 (22.2%) |
| 6 to 10 years | 1 (20.0%) | 0 (0%) | 1 (20.0%) | 2 (40.0%) | 2 (40.0%) |
| 11 to 15 years | 2 (40.0%) | 4 (80.0%) | 1 (20.0%) | 0 (0%) | 1 (20.0%) |
| 16 to 20 years | 1 (16.7%) | 0 (0%) | 3 (50.0%) | 3 (50.0%) | 1 (16.7%) |
| 21 years or more | 4 (66.7%) | 2 (33.3%) | 1 (16.7%) | 1 (16.7%) | 1 (16.7%) |
| Years involved in research | $p = 0.132^{F}$ | $p = 0.593^{C}$ | $p = 0.901^{C}$ | $p = 1.00^{F}$ | $p = 0.555^{F}$ |
| ≤10 years | 22 (22.7%) | 28 (28.9%) | 30 (30.9%) | 24 (24.7%) | 28 (28.9%) |
| >10 years | 7 (41.2%) | 6 (35.3%) | 5 (29.4%) | 4 (23.5%) | 3 (17.6%) |
| Job hours as clinician | $p = 0.887^{F}$ | $p = 0.107^{C}$ | $p = 0.578^{C}$ | $p = 0.840^{F}$ | $p = 0.506^{F}$ |
| Full time (≥ 35 hours/week) | 10 (25.0%) | 10 (25.0%) | 15 (37.5%) | 11 (27.5%) | 14 (35.0%) |
| Part time (< 35 hours/week) | 8 (29.6%) | 12 (44.4%) | 6 (22.2%) | 5 (18.5%) | 5 (18.56%) |
| Per diem (as needed) | 5 (27.8%) | 7 (38.9%) | 6 (33.3%) | 4 (22.2%) | 5 (27.8%) |
| No clinical practice involvement | 6 (20.7%) | 5 (17.2%) | 8 (27.6%) | 8 (27.6%) | 7 (24.1%) |
| Years of clinical experience | $p = 0.856^{F}$ | $p = 0.420^{F}$ | $p = 0.216^{F}$ | $p = 0.211^{F}$ | $p = 0.558^{F}$ |
| None | 1 (16.7%) | 1 (16.7%) | 1 (16.7%) | 0 (0%) | 1 (16.7%) |
| 1 to 5 years | 13 (28.9%) | 17 (37.8%) | 9 (20.0%) | 8 (17.8%) | 11 (24.4%) |
| 6 to 10 years | 4 (23.5%) | 4 (23.5%) | 7 (41.2%) | 6 (35.3%) | 4 (23.5%) |
| 11 to 15 years | 4 (30.8%) | 4 (30.8%) | 6 (46.2%) | 5 (38.5%) | 4 (30.8%) |
| 16 to 20 years | 3 (33.3%) | 4 (44.4%) | 2 (22.2%) | 1 (11.1%) | 1 (11.1%) |
| 21 years or more | 4 (16.7%) | 4 (16.7%) | 10 (41.7%) | 8 (33.3%) | 10 (41.7%) |
| Years of clinical experience | $p = 0.758^{C}$ | $p = 0.473^{C}$ | $p = 0.109^{C}$ | $p = 0.231^{C}$ | $p = 0.285^{C}$ |
| ≤10 years | 18 (26.5%) | 22 (32.4%) | 17 (25.0%) | 14 (20.6%) | 16 (23.5%) |
| >10 years | 11 (23.9%) | 12 (26.1%) | 18 (39.1%) | 14 (30.4%) | 15 (32.6%) |
| Highest degree (education level) | $p = 0.167^{F}$ | $p = 0.870^{F}$ | $p = 0.260^{F}$ | $p = 0.838^{F}$ | $p = 0.619^{F}$ |
| Baccalaureate Degree | 7 (18.9%) | 10 (27.0%) | 14 (37.8%) | 8 (21.6%) | 11 (29.7%) |
| Master’s Degree | 12 (22.6%) | 16 (30.2%) | 13 (24.5%) | 13 (24.5%) | 15 (28.3%) |
| Doctoral Degree | 6 (35.3%) | 5 (29.4%) | 4 (23.5%) | 5 (29.4%) | 4 (23.5%) |
| Master’s Degree in Progress | 1 (50.0%) | 1 (50.0%) | 1 (50.0%) | 0 (0%) | 1 (50.0%) |
| Doctoral Degree in Progress | 3 (60.0%) | 2 (40.0%) | 3 (60.0%) | 2 (40.0%) | 0 (0%) |

| | | | | | |
|---|---|---|---|---|---|
| Training in health research | p = 0.601[F] | p = 0.833[F] | p = 0.133[F] | p = 0.736[F] | p = 0.788[F] |
| No training | 9 (22.0%) | 11 (26.8%) | 17 (41.5%) | 9 (22.0%) | 12 (29.3%) |
| Undergraduate-level training | 2 (16.7%) | 4 (33.3%) | 4 (33.3%) | 2 (16.7%) | 2 (16.7%) |
| Graduate-level training | 18 (29.5%) | 19 (31.1%) | 14 (23.0%) | 17 (27.9%) | 17 (27.9%) |
| Training in EBP course | p = 0.638[C] | p = 0.358[C] | p = 0.933[C] | p = 0.483[C] | p = 0.061[C] |
| Yes | 12 (27.9%) | 15 (34.9%) | 13 (30.2%) | 9 (20.9%) | 16 (37.2%) |
| No | 17 (23.9%) | 19 (26.8%) | 22 (31.0%) | 19 (26.8%) | 15 (21.1%) |

Reported values are frequency (%) of correct responses, [C] chi-square test, [F] Fisher's Exact test

* ** significant values

### Appendix Table 2. Factors associated with perceived usefulness of five approaches

| **Variables** | **Approach** | | | | |
|---|---|---|---|---|---|
| | **RR/RRR** | **OR/ROR** | **RD/ARR** | **NNT** | **Risk in treatment vs. control groups** |
| Primary work setting | p = 0.720[A] | p = 0.707[A] | p = 0.871[A] | p = 0.679[A] | p = 0.401[A] |
| Clinical Nutrition | 4.67±1.60 | 4.51±1.53 | 4.38±1.80 | 4.18±1.89 | 4.48±1.64 |
| Social Services/Public Health | 5.14±1.21 | 5.29±1.25 | 5.14±1.57 | 5.43±1.13 | 5.29±1.11 |
| Primarily Research | 4.75±1.58 | 4.88±1.25 | 4.00±1.20 | 3.75±1.16 | 4.63±1.77 |
| Primarily Teaching | 5.00±1.15 | 4.86±1.21 | 5.00±1.41 | 4.29±1.70 | 5.14±1.07 |
| Both Research & Teaching | 5.38±0.92 | 4.75±1.39 | 5.13±0.83 | 4.38±2.00 | 5.13±1.13 |
| Government | 6.50±0.71 | 6.00±0.00 | 5.00±1.41 | 6.00±1.41 | 6.00±1.41 |
| Private Practice | 4.92±1.44 | 5.17±1.47 | 4.50±1.31 | 4.64±1.96 | 4.67±1.37 |
| Industry | 4.00±1.41 | 4.00±1.41 | 4.50±2.12 | 4.50±3.54 | 4.50±3.54 |
| Other | 4.57±1.99 | 5.00±2.31 | 4.71±2.87 | 4.14±1.95 | 3.43±2.30 |
| Primary work setting | p = 0.442[A] | p = 0.522[A] | p = 0.520[A] | p = 0.742[A] | **p = 0.035[A]** |
| Clinical practice | 4.71±1.57 | 4.62±1.53 | 4.40±1.72 | 4.25±1.90 | 4.51±1.59 |
| Non-clinical practice | 5.09±1.24 | 4.94±1.23 | 4.79±1.30 | 4.53±1.67 | **5.06±1.39*** |
| Other | 4.57±1.99 | 5.00±2.31 | 4.71±2.87 | 4.14±1.95 | **3.43±2.30**** |
| Years involved in research | p = 0.800[A] | p = 0.835[A] | p = 0.629[A] | p = 0.979[A] | p = 0.320[A] |
| None | 4.74±1.54 | 4.65±1.60 | 4.50±1.78 | 4.31±1.73 | 4.38±1.68 |
| 1 to 5 years | 5.15±1.49 | 4.85±1.51 | 4.52±1.58 | 4.31±2.22 | 4.70±1.64 |
| 6 to 10 years | 4.20±1.92 | 4.80±1.48 | 5.40±1.52 | 4.60±0.55 | 5.20±1.30 |
| 11 to 15 years | 4.80±1.30 | 4.60±1.52 | 4.80±1.30 | 4.25±2.36 | 4.60±1.34 |
| 16 to 20 years | 4.67±1.03 | 4.50±0.84 | 3.67±1.51 | 4.00±1.67 | 5.00±1.67 |
| 21 years or more | 4.83±1.60 | 5.50±0.84 | 5.00±1.79 | 4.83±1.94 | 5.83±0.75 |
| Years involved in research | p = 0.880[T] | p = 0.666[T] | p = 0.855[T] | p = 0.916[T] | p = 0.118[T] |
| ≤10 years | 4.82±1.55 | 4.71±1.56 | 4.55±1.71 | 4.32±1.83 | 4.51±1.65 |
| >10 years | 4.76±1.25 | 4.88±1.11 | 4.47±1.59 | 4.38±1.86 | 5.18±1.33 |
| Job hours as clinician | p = 0.305[A] | p = 0.676[A] | p = 0.798[A] | p = 0.870[A] | p = 0.411[A] |
| Full time (≥ 35 hours/week) | 4.78±1.67 | 4.60±1.53 | 4.50±1.97 | 4.38±1.94 | 4.65±1.73 |
| Part time (< 35 hours/week) | 4.44±1.63 | 4.59±1.65 | 4.35±1.55 | 4.07±1.80 | 4.19±1.63 |
| Per diem (as needed) | 4.83±1.38 | 4.83±1.58 | 4.50±1.47 | 4.47±1.84 | 4.61±1.38 |
| No clinical practice involvement | 5.21±1.15 | 5.00±1.28 | 4.79±1.54 | 4.41±1.74 | 4.93±1.58 |
| Years of clinical experience | p = 0.902[A] | p = 0.926[A] | p = 0.711[A] | p = 0.323[A] | p = 0.674[A] |
| None | 5.00±1.26 | 4.50±1.38 | 3.50±1.05 | 3.17±1.47 | 4.17±1.72 |
| 1 to 5 years | 4.93±1.36 | 4.67±1.37 | 4.69±1.59 | 4.09±1.81 | 4.62±1.57 |

| | | | | | |
|---|---|---|---|---|---|
| 6 to 10 years | 4.65±1.37 | 4.94±1.30 | 4.35±1.54 | 4.71±1.53 | 4.71±1.36 |
| 11 to 15 years | 5.08±1.61 | 5.08±1.44 | 4.54±1.81 | 4.15±1.46 | 4.00±1.35 |
| 16 to 20 years | 4.56±2.19 | 4.56±2.19 | 4.75±2.25 | 5.00±2.55 | 4.88±2.36 |
| 21 years or more | 4.63±1.66 | 4.67±1.74 | 4.58±1.86 | 4.63±1.93 | 4.88±1.75 |
| Years of clinical experience | $p = 0.672^T$ | $p = 0.894^T$ | $p = 0.759^T$ | $p = 0.257^T$ | $p = 0.951^T$ |
| ≤10 years | 4.87±1.34 | 4.72±1.34 | 4.50±1.56 | 4.17±1.74 | 4.60±1.52 |
| >10 years | 4.74±1.73 | 4.76±1.73 | 4.60±1.88 | 4.57±1.93 | 4.62±1.77 |
| Highest degree (education level) | $p = 0.239^A$ | $p = 0.062^A$ | $p = 0.710^A$ | $p = 0.528^A$ | $p = 0.119^A$ |
| Baccalaureate Degree | 4.51±1.69 | 4.24±1.72 | 4.39±1.92 | 4.19±1.84 | 4.17±1.76 |
| Master's Degree | 4.81±1.40 | 4.85±1.31 | 4.53±1.58 | 4.35±1.80 | 4.62±1.58 |
| Doctoral Degree | 5.41±1.18 | 5.06±1.25 | 4.59±1.54 | 4.24±1.75 | 5.29±1.05 |
| Master's Degree in Progress | 6.00±0.00 | 6.50±0.71 | 6.00±0.00 | 6.50±0.71 | 6.00±1.41 |
| Doctoral Degree in Progress | 4.60±1.95 | 5.40±1.82 | 5.00±2.00 | 4.60±2.51 | 4.80±1.92 |
| Training in health research | $p = 0.257^A$ | $p = 0.490^A$ | $p = 0.660^A$ | $p = 0.437^A$ | **$p = 0.027^A$** |
| No training | 4.56±1.61 | 4.51±1.70 | 4.35±1.94 | 4.03±1.91 | **4.20±1.74**** |
| Undergraduate-level training | 5.33±1.78 | 4.83±1.70 | 4.75±1.86 | 4.50±1.83 | 4.08±1.78 |
| Graduate-level training | 4.89±1.36 | 4.87±1.31 | 4.62±1.47 | 4.49±1.77 | **4.98±1.42*** |
| Training in EBP course | **$p = 0.004^T$** | $p = 0.063^T$ | **$p = 0.010^T$** | $p = 0.084^T$ | $p = 0.059^T$ |
| Yes | 5.33±1.25 | 5.05±1.17 | 5.02±1.35 | 4.71±1.77 | 4.98±1.44 |
| No | 4.51±1.57 | 4.55±1.65 | 4.24±1.81 | 4.10±1.83 | 4.39±1.69 |

Reported values are mean ± standard deviation, [A] one-way ANOVA, [T] Independent samples t-test

* ** significant values

### Appendix Table 3. Factors associated with confidence in five approaches

| **Variables** | **Approach** | | | | |
|---|---|---|---|---|---|
| | **RR/RRR** | **OR/ROR** | **RD/ARR** | **NNT** | **Risk in treatment vs. control groups** |
| Primary work setting | $p = 0.562^A$ | $p = 0.650^A$ | $p = 0.533^A$ | $p = 0.177^A$ | $p = 0.092^A$ |
| Clinical Nutrition | 4.07±1.46 | 4.07±1.50 | 4.13±1.72 | 3.59±1.50 | 3.84±1.53 |
| Social Services/Public Health | 4.83±0.98 | 5.00±1.73 | 5.57±1.13 | 5.29±0.95 | 5.43±1.13 |
| Primarily Research | 4.38±1.77 | 4.25±1.83 | 3.88±1.36 | 3.63±1.06 | 3.63±1.06 |
| Primarily Teaching | 4.29±0.95 | 4.43±0.79 | 4.00±1.63 | 3.86±1.35 | 4.14±1.57 |
| Both Research & Teaching | 5.25±1.75 | 5.13±1.46 | 5.13±1.55 | 4.50±1.85 | 5.38±1.41 |
| Government | 5.00±0.00 | 5.00±1.41 | 5.00±1.41 | 5.00±2.83 | 5.50±2.12 |
| Private Practice | 4.42±1.62 | 4.58±1.44 | 4.17±1.80 | 4.58±1.62 | 4.42±1.56 |
| Industry | 3.50±2.12 | 4.00±1.41 | 4.00±4.24 | 4.00±4.24 | 4.00±4.24 |
| Other | 3.86±2.19 | 4.14±2.41 | 4.43±2.64 | 3.86±2.48 | 4.00±2.45 |
| Primary work setting | $p = 0.214^A$ | $p = 0.255^A$ | $p = 0.416^A$ | $p = 0.248^A$ | $p = 0.108^A$ |
| Clinical practice | 4.12±1.48 | 4.15±1.50 | 4.14±1.72 | 3.75±1.55 | 3.93±1.54 |
| Non-clinical practice | 4.64±1.43 | 4.68±1.45 | 4.62±1.65 | 4.32±1.63 | 4.65±1.63 |
| Other | 3.86±2.19 | 4.14±2.41 | 4.43±2.64 | 3.86±2.48 | 4.00±2.45 |
| Years involved in research | $p = 0.471^A$ | $p = 0.159^A$ | $p = 0.546^A$ | $p = 0.871^A$ | $p = 0.221^A$ |
| None | 4.03±1.54 | 4.05±1.64 | 4.20±1.80 | 3.85±1.64 | 3.91±1.73 |

| | | | | | |
|---|---|---|---|---|---|
| 1 to 5 years | 4.41±1.47 | 4.33±1.47 | 4.44±1.72 | 3.93±1.80 | 4.37±1.42 |
| 6 to 10 years | 4.40±1.14 | 4.60±1.14 | 4.80±1.30 | 4.60±0.55 | 4.20±0.84 |
| 11 to 15 years | 4.40±2.30 | 4.80±1.10 | 3.60±1.67 | 4.00±1.73 | 4.20±2.05 |
| 16 to 20 years | 5.00±1.79 | 5.17±1.47 | 3.83±2.14 | 3.67±1.86 | 4.17±2.04 |
| 21 years or more | 5.00±0.00 | 5.50±0.84 | 5.33±1.51 | 4.50±1.64 | 5.67±0.82 |
| Years involved in research | p = 0.096[T] | **p = 0.012[T]** | p = 0.992[T] | p = 0.727[T] | p = 0.131[T] |
| ≤10 years | 4.16±1.50 | 4.15±1.57 | 4.30±1.75 | 3.91±1.65 | 4.05±1.62 |
| >10 years | 4.82±1.55 | 5.18±1.13 | 4.29±1.86 | 4.06±1.68 | 4.71±1.76 |
| Job hours as clinician | p = 0.790[A] | p = 0.220[A] | p = 0.846[A] | p = 0.411[A] | p = 0.252[A] |
| Full time (≥ 35 hours/week) | 4.10±1.39 | 3.98±1.46 | 4.10±1.81 | 3.73±1.60 | 3.98±1.56 |
| Part time (< 35 hours/week) | 4.19±1.60 | 4.19±1.69 | 4.41±1.76 | 3.70±1.68 | 3.78±1.48 |
| Per diem (as needed) | 4.39±1.75 | 4.67±1.57 | 4.33±1.50 | 4.11±1.49 | 4.56±1.62 |
| No clinical practice involvement | 4.45±1.53 | 4.66±1.49 | 4.45±1.90 | 4.31±1.75 | 4.48±1.88 |
| Years of clinical experience | p = 0.278[A] | p = 0.770[A] | p = 0.500[A] | p = 0.139[A] | p = 0.368[A] |
| None | 3.83±1.60 | 3.83±1.94 | 3.00±1.55 | 3.00±1.26 | 3.17±1.47 |
| 1 to 5 years | 4.44±1.49 | 4.29±1.47 | 4.22±1.66 | 3.82±1.67 | 4.33±1.67 |
| 6 to 10 years | 4.53±1.50 | 4.47±1.50 | 4.71±1.86 | 4.76±1.44 | 4.47±1.37 |
| 11 to 15 years | 4.67±1.78 | 4.77±1.54 | 4.38±1.85 | 4.08±1.71 | 3.54±1.51 |
| 16 to 20 years | 3.44±1.33 | 3.89±1.17 | 4.33±2.00 | 3.22±1.56 | 3.89±1.27 |
| 21 years or more | 3.92±1.47 | 4.25±1.82 | 4.42±1.79 | 3.96±1.68 | 4.25±1.96 |
| Years of clinical experience | p = 0.184[T] | p = 0.915[T] | p = 0.644[T] | p = 0.663[T] | p = 0.365[T] |
| ≤10 years | 4.41±1.49 | 4.29±1.51 | 4.24±1.74 | 3.99±1.64 | 4.26±1.60 |
| >10 years | 4.02±1.56 | 4.33±1.63 | 4.39±1.81 | 3.85±1.66 | 3.98±1.72 |
| Highest degree (education level) | **p < 0.001[A]** | **p < 0.001[A]** | p = 0.359[A] | p = 0.082[A] | p = 0.073[A] |
| Baccalaureate Degree | **3.51±1.59*** | **3.57±1.69*** | 3.39±2.09 | 3.32±1.68 | 3.57±1.74 |
| Master's Degree | 4.40±1.34 | 4.42±1.34 | 4.38±1.47 | 4.19±1.51 | 4.30±1.56 |
| Doctoral Degree | **5.35±1.22**** | **5.35±1.11**** | 4.53±1.55 | 4.12±1.62 | 4.71±1.53 |
| Master's Degree in Progress | 5.00±0.00 | 6.00±1.41 | 6.00±1.41 | 4.50±0.71 | 5.50±0.71 |
| Doctoral Degree in Progress | 4.40±1.52 | 4.40±1.52 | 4.80±2.59 | 4.80±2.28 | 4.40±1.52 |
| Training in health research | **p < 0.001[A]** | **p = 0.002[A]** | p = 0.086[A] | **p = 0.001[A]** | **p = 0.002[A]** |
| No training | **3.38±1.61**** | **3.63±1.73**** | 3.83±1.99 | **3.20±1.58**** | **3.44±1.72**** |
| Undergraduate-level training | **4.92±1.24*** | **4.83±1.03*** | 4.83±1.80 | 4.33±1.67 | **4.75±1.36*** |
| Graduate-level training | **4.70±1.24*** | **4.66±1.36*** | 4.51±1.53 | **4.34±1.53*** | **4.51±1.50*** |
| Training in EBP course | **p < 0.001[T]** | **p = 0.036[T]** | **p = 0.014[T]** | **p = 0.034[T]** | **p = 0.007[T]** |
| Yes | 4.91±1.27 | 4.67±1.29 | 4.81±1.55 | 4.35±1.59 | 4.67±1.49 |
| No | 3.86±1.53 | 4.08±1.66 | 3.99±1.82 | 3.68±1.64 | 3.83±1.66 |

Reported values are mean ± standard deviation, [A] one-way ANOVA, [T] Independent samples t-test

* ** significant values
